# Metabolome-defined obesity and the risk of future diabetes and mortality

**DOI:** 10.1101/2021.11.03.21265744

**Authors:** Filip Ottosson, Einar Smith, Ulrika Ericson, Salvatore Di Somma, Paola Antonini, Peter M Nilsson, Céline Fernandez, Olle Melander

**Author notes:** Corresponding author: Filip Ottosson.

## Abstract

**Background:** Obesity is a key risk factor for type 2 diabetes, however, up to 20% of patients are normal weight. Our aim was to identify metabolite patterns reproducibly predictive of BMI, and subsequently to test if lean individuals who carry an obese metabolome are at hidden high risk of obesity related diseases, such as diabetes.

**Methods:** We measured 109 metabolites in fasted plasma samples of 7663 individuals from two Swedish and one Italian population-based cohort. Ridge regression models were used to predict BMI using the plasma metabolites. Individuals with a predicted BMI either more than 5 kg/m^2^ higher (overestimated) or lower (underestimated) than their actual BMI were characterized as outliers and further investigated for obesity related risk factors and future risk of diabetes and mortality.

**Results:** The plasma metabolome could predict BMI in all cohorts (r^2^ = 0.48, 0.26 and 0.19). The overestimated group had a BMI similar to individuals correctly predicted as normal weight, similar waist circumference, were not more likely to change weight over time but had a 2 times higher risk of future diabetes and an 80 % increased risk of all-cause mortality. These associations remained after adjustments for obesity-related risk factors and lifestyle parameters.

**Conclusions:** We found that lean individuals with an obese metabolome, have an increased risk for diabetes and all-cause mortality compared to lean individuals with a healthy metabolome. Metabolomics may be used to identify hidden high-risk individuals, in order to initiate lifestyle and pharmacological interventions.

## Introduction

The epidemic of obesity is a global health burden resulting in 2.8 million deaths each year^1^ co-occurring with a rapid increased prevalence of diabetes. Even if obesity is the key modifiable risk factor for diabetes, up to 20% of type 2 diabetes patients are normal weight^2^. Obesity is imprecisely defined by body-mass index (BMI), a heterogeneous definition that may not accurately describe the associations between obesity and its comorbidities^3^. In this light, there are individuals with hidden increased risk for obesity-related health issues despite having a normal BMI. Conversely, some individuals may have a high BMI but remain more resistant to common co-occurring pathologies, frequently referred to as metabolically healthy obesity (MHO)^4^.

The recent decade, metabolomics has appeared as a useful discipline in characterizing the human metabolism^5^. Several studies have found associations between the plasma metabolome and obesity^6, 7^, highlighting potential obesity related metabolic dysregulation. These metabolites include among others amino acids and metabolites of amino acid catabolism, lipids and nucleotides. Several of these plasma metabolites have also shown BMI independent associations with future risk of diabetes^8-11^, cardiovascular disease^12-14^ and mortality^15, 16^ in several prospective studies. This indicates that these metabolites may have the potential to refine the definition of obesity beyond anthropometric measurements. In a study conducted by Cirulli et al, a pattern of 49 plasma metabolites was strongly associated with BMI and was applied to classify individuals as obese or non-obese^17^. Interestingly, individuals who were classified as obese according to the metabolome had twice as high risk for future cardiovascular events and more pronounced insulin resistance compared to BMI-matched individuals who were classified as non-obese by the metabolome^17^. This indicates that a portion of the normal weight population is at an increased risk of obesity related diseases, despite being characterized as lean according to anthropometric measurements.

We applied plasma metabolomics in 7663 individuals from three population-based cohorts, in order to robustly identify two separate strata of the population: normal weight individuals with an obese metabolome and obese individuals with a normal weight metabolome. Hypothesizing that these metabolome alterations associate with obesity-related pathologies, we investigated whether the risk of future diabetes and all-cause mortality was different in these strata compared to their normal weight and obese counterparts.

## Methods

### Study Samples

Plasma levels of metabolites were measured in 7663 individuals from two Swedish and one Italian population-based cohort. This included two cross-sectional cohorts, the Malmö Offspring Study (MOS) and the Cilento in Aging Outcomes Study (CIAO) and the prospective cohort, the Malmö Diet and Cancer Study (MDC). The ethics committee of Lund University approved the study protocols for MOS (DNR 2012/594) and MDC (DNR 2009/633), The Regional Board of Ethics ASL Napoli Sud (20171220) approved the protocols for CIAO and all participants provided written informed consent. Participants who were diagnosed with diabetes or a BMI lower than 18 kg/m^2^ were excluded from the analyses. An overview of the three cohorts, including excluded participants can be found in Figure S1.

The Malmö Diet and Cancer Study (MDC) is a population-based prospective cohort consisting of 28 449 individuals. The cardiovascular cohort of the MDC was designed to study the epidemiology of carotid artery disease with participants being enrolled between 1991 and 1996^18^. Among the 5405 participants who came fasted, citrate plasma was obtained from 3833 participants for metabolite analysis. During an average follow-up time of 18.2 years 491 participants developed T2DM and within 19.7 years, 967 individuals died. Participants from the cardiovascular cohort of MDC were further invited to a follow-up examination between 2007 and 2012, where clinical examinations were performed.

MOS is an ongoing population-based cohort study where adult (>18 years old) children and grand-children from the MDC study are recruited^19^. Participants were invited via letter and visited the research clinic where over-night fasting EDTA plasma was collected and anthropometric measurements were performed. Levels of metabolites were measured in all participants with available plasma samples (N=3430).

CIAO is a population-based cohort, based in the area of Cilento in the Campania region in South Italy. A random sample of middle-age (50-67 years) individuals from Cilento were invited via the local primary health care^20^, with a participation rate of 55 %. A total of 821 individuals had over-night fasted EDTA plasma samples available for metabolite analysis.

### Endpoint definitions, biochemical measurements and lifestyle assessments

Endpoint retrieval was performed through record linkage of the personal identification number of each Swedish citizen with Swedish local or national registries^21^. T2D was defined as a fasting plasma glucose of >7.0 mmol/L or a history of physician diagnosis of T2D or being on antidiabetic medication or having been registered in local or national Swedish diabetes registries^22^. Details about biochemical measurements, methods for assessment of dietary intakes and physical activity are found in Material S1.

### Analytical Procedure

Profiling of plasma metabolites was performed using a UPLC-QTOF-MS System (Agilent Technologies 1290 LC, 6550 MS, Santa Clara, CA, USA) and has previously been described in detail^23^. Briefly, plasma samples stored at -80 □C were thawed and extracted by addition of six volumes of extraction solution. The extraction solution consisted of 80:20 methanol/water. Extracted samples were separated on an Acquity UPLC BEH Amide column (1.7 *µ*m, 2.1 * 100mm; Waters Corporation, Milford, MA, USA). Information about quality control, normalization and metabolite annotation is found in Material S1.

### Statistical analysis

In each cohort, metabolite levels were mean centred and unit variance scaled. Outliers were defined as more than 5 standard deviation (sd) units away from the mean and were imputed as either -5 or 5. Missing values were imputed as -5. BMI was modelled using ridge regression with the R package *glmnet* (v 3.0-2)^24^. The λ-parameter was optimized using *cv*.*glmnet*, minimizing the mean-squared error, varying λ between 1000 and 0.01. Cross-validated mean-squared error for different λ are found in Figure S2. Model training was performed in 80% randomly selected participants from MOS and validation was performed in the remaining 20%. The model was replicated in MDC and CIAO. Prediction of obesity and overweight was assessed using the area under the receiver operating characteristics curve (AUC). BMI was mean centred and unit variance scaled for the ridge regression modelling. The levels of all 108 metabolite levels were compared between BMI-classes using ANOVA and Tukey’s post hoc test. In MDC, the association between BMI-class and longitudinal weight-change was analysed using ANOVA, while the association between BMI-class and incident diabetes and all-cause mortality was analysed using Cox regression models. Model 1 was adjusted for age and sex, model 2 additionally adjusted for fasting levels of glucose, triglyceride and HDL cholesterol and for smoking status. Differences in dietary intake and physical activity between BMI-classes were analysed using ANOVA and linear regression. All statistical analyses were performed in R.3.6.1. ANOVA was performed using *aov*, Tukey’s post hoc tests using *TukeysHSD*, AUC using *pROC*, Cox regression using *survival* and data visualizations using *ggplot2*. Scripts are publicly available at: https://github.com/immu-flo/metabolome_BMI.

## Results

Metabolomic measurements were performed in three population-based cohorts, MOS, MDC and CIAO. There were several differences between the three cohorts, most notably MOS consisted of participants from the age of 18-70, while the participants in both MDC and CIAO were middle age (50-65 years old). There were also differences in several metabolic risk factors between the cohorts, such as BMI and fasting glucose levels (Table 1).

**Table 1.** Characteristics of the participants in the Malmö Diet and Cancer Study (MDC), Malmö Offspring Study (MOS) and Cilento in Aging Outcomes Study (CIAO).

|  | **MOS (N=3263)** | **MDC (N=3579)** | **CIAO (N=821)** | **p** |
| --- | --- | --- | --- | --- |
| **Age (years)** | 41.59 (14.11) | 57.59 (6.00) | 57.76 (4.53) | <0.001 |
| **Sex (% female)** | 1709 (52.4) | 2122 (59.3) | 466 (56.8) | <0.001 |
| **BMI (kg/m<sup>2</sup>)** | 26.02 (4.63) | 25.56 (3.72) | 28.08 (5.39) | <0.001 |
| **Waist (cm)** | 89.94 (13.69) | 82.97 (12.32) | 96.46 (69.58) | <0.001 |
| **Glucose (mmol/l)** | 5.32 (0.57) | 4.96 (0.52) | 5.58 (0.65) | <0.001 |
| **HDL (mmol/l)** | 1.62 (0.48) | 1.40 (0.37) | 1.56 (0.39) | <0.001 |
| **TG (mmol/l)** | 1.12 (0.67) | 1.28 (0.61) | 1.35 (0.78) | <0.001 |
| **Current Smoker (%)** | 457 (14.0) | 956 (27.5) | 0 (0.0) | <0.001 |

### A wide range of metabolites associates with obesity

We used 109 plasma metabolites to find associations with BMI in the 7663 participants from the three investigated cohorts. A ridge regression model was trained in a subset consisting of 80 % randomly selected participants from MOS (N=2611) (BMI = 26.0 ±4.6). In the validation set, consisting of the remaining 20 % (N=652) (BMI = 26.0 ±4.7), the ridge regression model could explain almost half of the variation of BMI (r^2^=0.476) (Figure 1A). The model was able to predict both obesity (AUC=0.936) and overweight (AUC=0.803) compared to normal weight. In both external validation cohorts, MDC and CIAO, the ridge regression model was able to predict BMI (MDC: r^2^=0.256, CIAO: r^2^=0.196) (Figure 1B-C). In both MDC and CIAO, the model could predict obesity (AUC=0.86 and AUC=0.86) and overweight (AUC =0.71 and AUC=0.70) compared to normal weight. The BMI predicted by the ridge regression model is further referred to as metabolic BMI.

**Figure 1.**
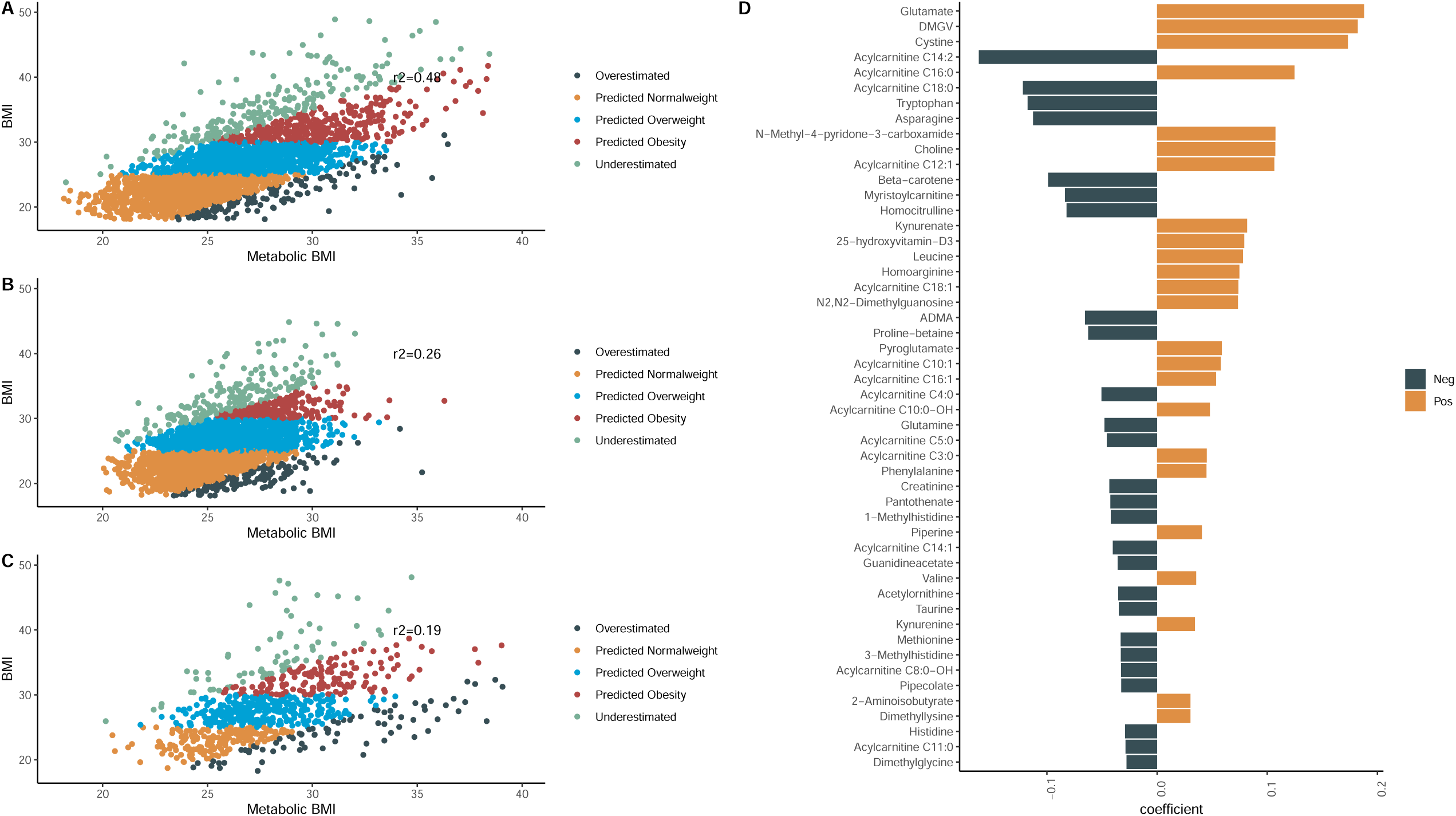
Metabolite Prediction of BMI. Correlation between ridge regression-based prediction of BMI (metabolic BMI) and BMI in Malmö Offspring Study (N=3263) (A), Malmö Diet and Cancer Study (N=3579) (B) and the Cilento in Aging Outcomes study (N=821) (C). The outlier groups were defined as having a metabolic BMI > 5 kg/m^2^ higher than BMI (Overestimated) or a metabolic BMI < 5 kg/m^2^ lower than BMI (Underestimated). The remaining participants were categorized according to their BMI as normal weight (BMI<25 kg/m^2^), overweight (BMI= 25-30 kg/m^2^) and obesity (BMI>30 kg/m^2^). The coefficients of the top 30 metabolites in the ridge regression model is shown in (D).

Metabolites of several different biochemical classes contributed to the obesity-predictive model, such as amino acids (glutamate, cystine and tryptophan), acylcarnitines (C14:2-carnitine, C18:0-carnitine and 16:0-carnitine), nucleotides (N2,N2-dimethylguanosine) and food derived metabolites (beta-carotene and proline betaine) (Figure 1D). The metabolites that contributed most to the model in positive direction were dimethylguanidino valerate (DMGV) and glutamate, and in the negative direction C14:2-carnitine and C18:2-carnitine.

### Outliers of metabolome-predicted BMI have different levels of circulating lipids

Next, individuals were classified into five different groups according to their BMI and metabolomic BMI. Participants whose BMI could be predicted within an error margin of 5 kg/m^2^ (metabolomic BMI-BMI: -5kg/m^2^-5kg/m^2^) were characterized according to their BMI as normal weight (NW) (BMI<25 kg/m^2^), overweight (OW) (BMI=25-30 kg/m^2^) or obese (OB) (BMI>30kg/m^2^). Participants whose BMI were not predicted within the error margin were either classified as overestimated (OE) (metabolomic BMI-BMI > 5kg/m^2^) or underestimated (UE) (metabolomic BMI-BMI < -5kg/m^2^).

In MOS, 88.7 % (N=2894) had a BMI predicted within the error margin, 4.1 % were classified as OE (N=133) and 7.2 % as UE (N=236). Similar proportion of participants were predicted within the error margin (90.1 %) in MDC, while 3.8 % were characterized as OE (N=132) and 6.1 % as UE (N=210). In CIAO, the proportion of well-predicted participants were slightly lower, at 81.6 %. Among the remaining outliers, 9.7 % of the participants were OE (N=80) and 8.6 % were UE (N=71).

The average BMI in the OE groups (MOS=21.8, MDC=20.8 and CIAO=24.9) were similar to NW in all three cohorts, while the average BMI of the UE groups (MOS=35.4, MDC=33.6 and CIAO=37.9) were similar to OB. Despite this large difference in BMI between OE and UE, the metabolic BMI in OE (MOS=27.0, MDC=28.1 and CIAO=32.0) and UE (MOS=27.7, MDC=26.2 and CIAO=28.2) were similar to each other and were more similar to OW or OB (Figure 2A-F). There were no differences found in waist circumference between OE and NW in neither MOS (p=0.49) nor MDC (p=0.17). In CIAO, the average waist circumference in OE was 8 cm larger than in NW (p<0.001). Comparing the waist circumference of UE and OB, there were no differences in MDC (p=0.83) and in both MOS (difference= 2.6 cm, p=0.003) and CIAO (difference= 5cm, p=0.003) the average waist circumference in UE was slightly larger than in OB (Figure S3).

**Figure 2.**
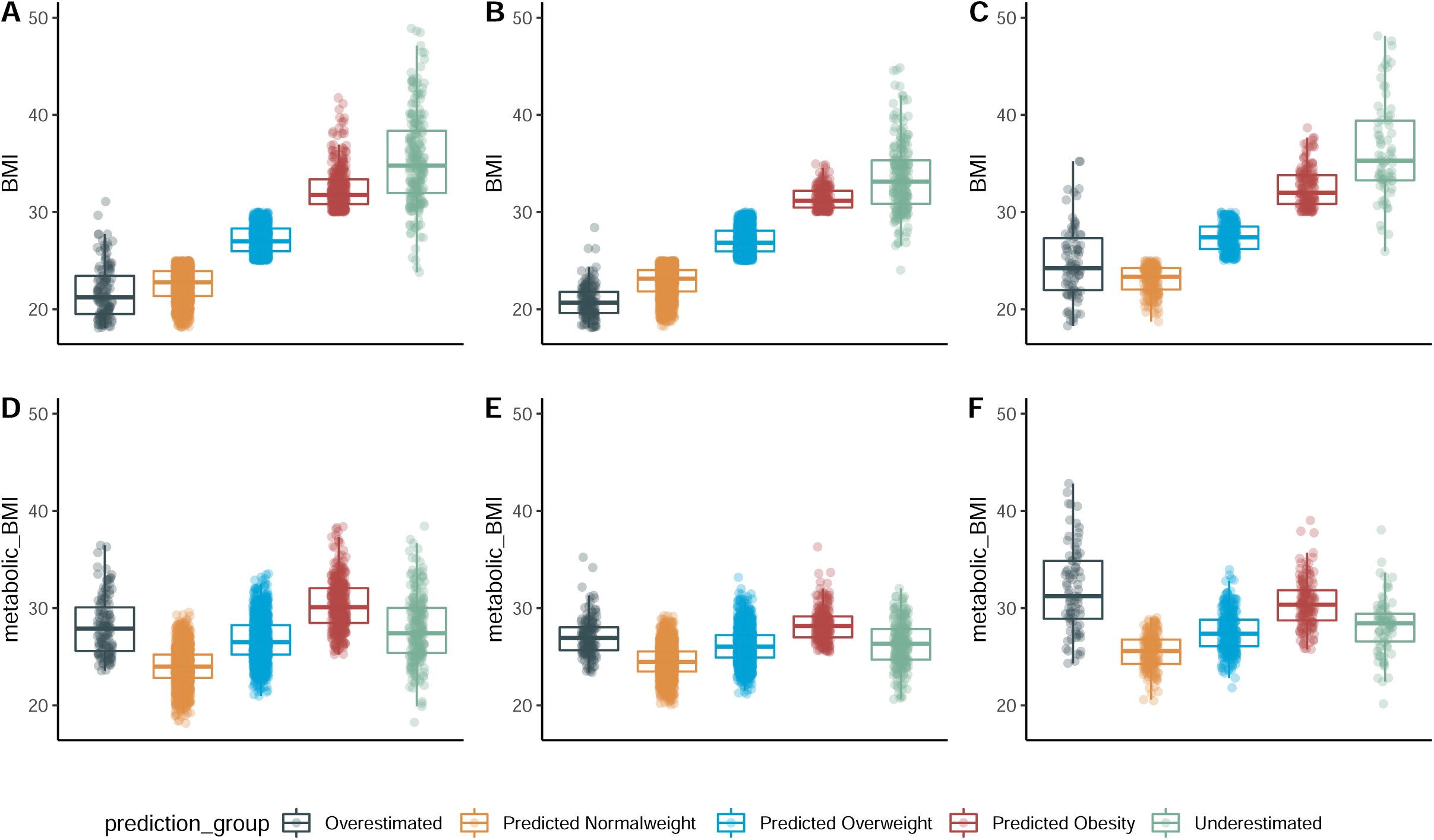
Differences in BMI (kg/m^2^) (A-C) and metabolic BMI (kg/m^2^) (D-F) between metabolic BMI outlier groups. Results from the Malmö Offspring Study (N=3263) are shown in (A) and (D), the Malmö Diet and Cancer Study (N=3579) in (B) and (E) and the Cilento in Aging Outcomes study (N=821) in (C and F).

Plasma levels of triglycerides were higher in OE compared to NW and lower in UE compared to OB in all three investigated cohorts (Figure S3). Plasma levels of HDL cholesterol were lower in OE compared to NW and higher in UE compared to OB in both MOS and CIAO, but no differences were observed in MDC (Figure S4). For fasting glucose, no differences were seen between OE and NW in any cohort, and slightly lower levels of glucose in UE compared to OB was only seen in CIAO (Figure S4). In MOS and MDC, OE had higher proportions of smokers compared to NW (p_diff_<0.001). The proportion of smokers in OE were 24.6, 47.2 and 26.0 %, compared to 13.3, 30.9 and 24.7 % in NW in MOS, MDC and CIAO respectively. All group-wise differences in metabolic risk factors are found in Table S1-S3.

### Metabolite level differences in outliers of metabolic obesity are consistent in independent cohorts

In MOS, the levels of 13 metabolites were significantly different (p<4.6e-4) between participants in OE and NW. These included several of the metabolites that strongly influenced metabolic BMI, such as DMGV, glutamate and beta-carotene. All thirteen metabolites had at least nominally significant differences, in consistent direction, between NW and OE in at least two out of three investigated cohorts. For five metabolites, DMGV, glutamate, beta-carotene, isoleucine and kynurenine, significant (p<0.05) differences were found in all cohorts (Figure S5). All group-wise differences in metabolite levels are found in Table S4.

### Lean individuals with metabolic obesity are at higher risk of future diabetes and premature death

Next, we wanted to investigate whether being an outlier of metabolic obesity is associated with risk of future obesity, diabetes and death. Prospective follow-up of obesity, diabetes and mortality was available in MDC but not in CIAO and MOS. During an average follow-up of 15.6 years, there were no difference in weight change between any of the five investigated groups (Figure S6) (Table S5). More specifically, there were no significant difference in weight change between baseline and follow-up between OE and NW (p=0.98). However, during an average follow-up time of 18.5 years, participants in OE had more than 2-fold higher risk of developing type 2 diabetes (HR per sd= 2.22, C.I=1.38-3.56, P=0.001) compared to NW. This association was attenuated but remained significant when adjusting for smoking status and fasting levels of glucose, triglycerides, HDL cholesterol. In the adjusted model, there were no significant difference in risk of future diabetes between OE and any of the other groups (Figure 3) (Table S6).

**Figure 3.**
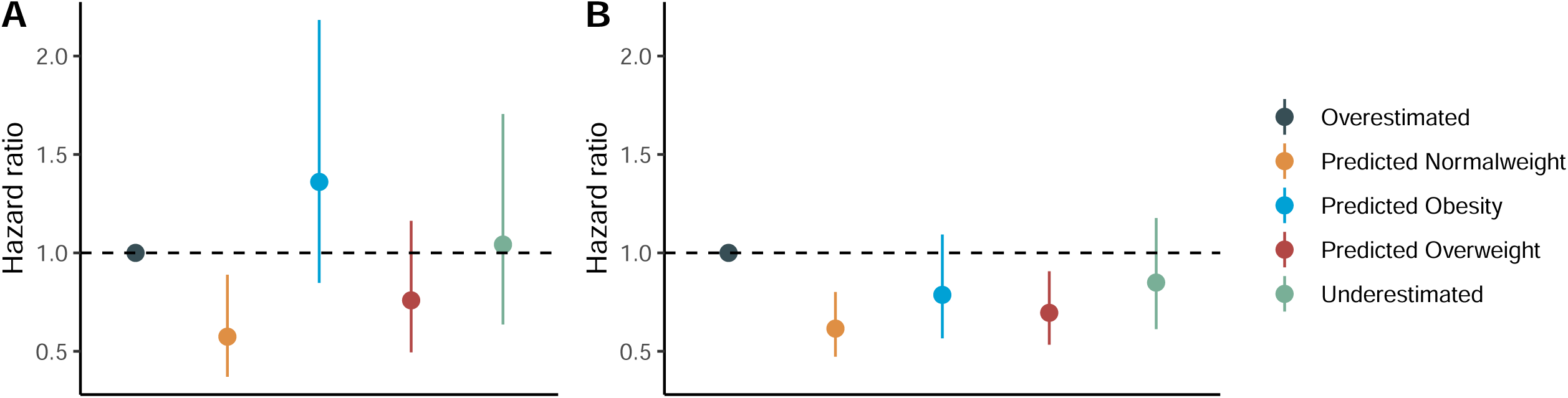
Associations between outlier groups of metabolic BMI and future risk of diabetes (A) and mortality (B) in the Malmö Diet and Cancer Study (N=3579). The hazard ratios (HR) are expressed as compared to the reference group (Overestimated). Error-bars correspond to 95 % confidence intervals. HR are calculated from Cox proportional hazard models, adjusted for age, sex, smoking status and fasting levels of glucose, HDL cholesterol and triglycerides.

Participants in OE compared to NW were also at almost twice as high risk of all-cause mortality within an average follow-up time of 19.7 years (HR=1.85, C.I=1.50-2.05, p<0.001). Interestingly, OE were also associated with an increased risk of all-cause mortality compared to OW (HR=1.72, C.I=1.30-2.29, p=0.0002) and OB (HR=1.56, C.I=1.09-2.24, p=0.015). After additional adjustments for smoking status and levels of glucose, triglycerides and HDL cholesterol, these associations were attenuated, but remained statistically significant regarding OE vs NW and OW (Figure 3) (Table S6). Participants in UE had a 53 % decreased risk of T2D compared to OB (HR=1.53, C.I=1.08-2.20, P=0.017), but were not at lower risk of all-cause mortality (HR=0.84, C.I=0.60-1.18, P=0.32). The association with T2D was attenuated after adjustments for potential confounders (HR=1.31, C.I=0.91-1.87, P=0.14) (Table S6).

### Life-style factors associated with metabolic obesity

Utilizing dietary assessments in MOS (N=1526) and MDC (N=3471), we tested whether seven dietary intakes differed between the five BMI-prediction groups. In MDC, the diet of OE consisted of significantly (p<0.05) more saturated fat and less fruits and vegetables (Figure S7), and whole-grain compared to NW. No differences in intakes were seen for polyunsaturated fats, fish and shellfish, meat or added sugar. UE consumed higher amounts of fruit and vegetables and lower amounts of polyunsaturated fats compared to OB (Table S7). In MOS, OE consumed significantly (p<0.05) lower amounts of fruits and vegetables compared to NW (Figure S7), but no differences were seen for any of the other examined dietary factors. The differences in intakes of fruits and vegetables between NW and OE were significant after adjustments for age, sex and smoking status in both MDC (beta=6.6, C.I=2.9-6.3, P=4.4e-4) and MOS (beta=8.9, C.I=3.6-14.1, P=1.0e-3). No differences were seen between UE and OB in dietary intakes in MOS (Table S7). Self-assessed levels of leisure-time physical activity were significantly (p<0.05) different between OE and NW but not between UE and OB in MOS, but no significant differences were seen in MDC (Table S8). In sensitivity analysis in MDC (N=3472), the risk for diabetes and all-cause mortality in OE compared to NW was only marginally changed after adjustments for intakes of fruits and vegetables, whole-grain, saturated fats and leisure-time physical activity (Table S9).

## Discussion

Utilizing the metabolome of 7663 individuals, the present study identifies a metabolite fingerprint of BMI (metabolic BMI) and investigates the risk of diabetes and mortality in individuals whose metabolic BMI differ from their actual BMI. Middle-aged individuals who are normal weight but have a metabolic BMI more than 5 kg/m^2^ above their actual BMI have a doubled risk of future diabetes and all-cause mortality compared to individuals who are normal weight both according to BMI and metabolic BMI. Similarly, individuals who are obese, but have a substantially lower metabolic BMI have decreased risk of diabetes compared to individuals who are obese both according to BMI and metabolic BMI. This study attempts to refine the definition of obesity by identifying strata of the population who, based on their metabolome, have different risk of diabetes and death compared to what their BMI indicates.

### Plasma metabolites are strongly associated with obesity across different cohorts

The association between the metabolome and obesity has been described in detail in several population-based cohorts^6, 17, 25, 26^. Consequently, most of the metabolites strongly influencing metabolic BMI in this study have documented associations with BMI or waist circumference in previous publications. For instance, these include DMGV^27, 28^, glutamate^29^ and the branched-chain amino acids (BCAA) leucine and valine^25^. Moreover, all the aforementioned metabolites have been associated with increased risk of future diabetes^10, 11, 14, 27, 30^. The associations between metabolites and BMI were relatively consistent over the three investigated cohorts. Metabolic BMI explained 47 % of the variation of BMI in the validation set of MOS. Despite the model being trained in MOS, it was able to explain a large portion of the variation of BMI (19-26 %) also in MDC and CIAO, although to a smaller degree than in MOS. This indicates that the BMI prediction model is not specific to the population of MOS, but rather is generalizable to independent cohorts with different characteristics.

The present study was conducted using data from a relatively limited number of 109 metabolites. Applying similar strategies using a larger number of metabolites from a wide range of biochemical classes should improve the predictive ability. For instance, in the study by Cirulli et al, extending a model utilising the 49 metabolites with strongest associations with BMI to a model containing 650 metabolites, improved the explained variation of BMI from 39 to 49 %^17^. Among the 49 metabolites, several were also important predictors of BMI in this study. Most notably, these included glutamate, asparagine, leucine, N2,N2-dimethylguanosine and kynurenate. It is noteworthy that although there are similarities between the studies, among the 49 metabolites, only ten were measured here. The majority of the remaining 39 metabolites were lipids that were not covered by the methodology applied in this study. Although it is reasonable to expect that a larger number of metabolites should result in an improved prediction, the explained variation of BMI in the present study ranged from 19-47 %, which is well in line with previous studies, ranging from 23-49 %^7, 17^.

### Outliers of metabolic BMI have an increased risk for diabetes and mortality

A subset of the participants (4.1-9.7 %) were classified as OE (metabolic BMI at least 5 kg/m^2^ above their actual BMI). The risk for future T2D was twice as high in OE compared to NW, despite having slightly lower average BMI. In line with previous findings^17^, individuals in OE compared to NW, had higher levels of triglycerides and lower levels of HDL cholesterol. Fasting glucose levels were however not significantly different between the groups. There were still significant differences in diabetes risk between OE and NW after adjustments for these potential confounders, although the association was slightly attenuated. Similarly, the risk of all-cause mortality was 80 % higher in OE compared to NW, an association that also remained significant after adjustments for potential confounders. We argue that characterizing this subset of the population is clinically relevant for two separate reasons. Firstly, this subset of the population is of high risk of diabetes and premature mortality but may likely be missed by conventional methods because of their low BMI and non-elevated fasting glucose levels. This is further stressed by our finding that OE do not differ in weight gain over time compared to NW. Thus, the metabolic BMI could help pinpoint this hidden high-risk subset of the population, motivating lifestyle and pharmacological interventions. Importantly, since metabolic BMI can identify increased risk of diabetes and mortality up to 20 years before the event, intervention strategies can be implemented early enough to potentially reach a substantial risk reduction. Secondly, alterations in metabolite levels in OE compared to NW could highlight metabolic pathways involved in the pathological process of diabetes. In the present study, the five metabolites that consistently differ between OE and NW; i.e. DMGV^27, 30^, glutamate^31, 32^, beta-carotene^10^, isoleucine^11, 32^ and kynurenine, have all been associated with future risk of type 2 diabetes in previous studies. If a causal link between the metabolites and either diabetes or mortality risk can be proven, such metabolites could be potential pharmacological target molecules. This strategy could be particularly efficient for OE since weight reduction per se is unlikely to be effective.

Our results suggest that differences between OE and NW in terms of risk of T2D and mortality may be partly explained by lifestyle factors. Individuals in OE consumed significantly less fruit and vegetables while having a higher prevalence of smoking. These factors themselves do not explain the difference in disease risk between OE and NW, since the associations with T2D and mortality remained significant after adjustments. However, given that life-style factors are strongly correlated with each other, it is possible that the differences between NW and OE in smoking status and dietary intakes of fruits and vegetables may be explained by a generally unhealthy lifestyle, typical for the OE individuals.

### The metabolome may identify metabolically healthy obesity

The proportion of participants characterized as UE (metabolic BMI at least 5 kg/m^2^ below their actual BMI) was between 6.1-8.6 % in the three cohorts. Similar to the comparison between OE and NW, the risk of developing diabetes was 50 % higher in OB compared to UE, despite the average BMI was 2-5 kg/m^2^ higher in UE. This association was however attenuated after adjustments for confounders. The phenomenon that a portion of the obese population may be protected from obesity related pathologies, commonly referred to as MHO, have been discussed extensively elsewhere and several definitions have been suggested^4, 33^. Our study suggests that defining metabolically healthy obesity based on metabolite levels may be an alternative. The levels of several T2D-related metabolites, such as DMGV, isoleucine and leucine were different between UE and OB. This is consistent with lower risk of diabetes in MHO^34^ and suggests that these metabolites may help characterize the metabolic difference between MHO and metabolically unhealthy obesity and possibly explain their different clinical prospects.

### Strengths and limitations

This study has several limitations. Firstly, although we show that the association between the metabolome and obesity is consistent across different cohorts from different geographical regions, major hurdles remain to reach clinical utility, including absolute quantification of metabolite levels, which is needed to determine the metabolic BMI clinically. Secondly, the outlier classifications were arbitrarily determined as a metabolic BMI 5 kg/m^2^ above or below the actual BMI and may not be the most clinically relevant classification. Thirdly, since metabolites only were measured at one time point, we could not provide any data regarding the stability of the metabolic BMI classifications over time. Finally, prospective analyses could only be performed in one out of three investigated cohorts, which calls for replication of the association between metabolic BMI and diabetes and mortality in other populations to confirm its validity.

## Data Availability

All data produced in the present study are available upon reasonable request to the authors

## Acknowledgements

The authors thank the participants in The Malmö Offspring Study, The Malmö Diet and Cancer Study and The Cilento on Ageing Outcomes Study for making this research possible.

## Supplemental Figure Legends

**Figure S1**. Study design flow chart, including exclusion criteria for all three studies: Malmö Offspring Study (MOS), Malmö Diet and Cancer Study (MDC) and Cilento on Ageing Outcomes Study (CIAO).

**Figure S2**. Optimization of λ in cross-validated Ridge-regression models.

**Figure S3**. Differences in waist circumference (cm) (A-C) and fasting plasma triglyceride levels (TG) (mmol/L) (D-F) between metabolic BMI outlier groups. Results from the Malmö Offspring Study (N=3263) are shown in (A) and (D), the Malmö Diet and Cancer Study (N=3579) in (B) and (E) and the Cilento in Aging Outcomes study (N=821) in (C and F).

**Figure S4**. Differences in fasting plasma HDL cholesterol levels (mmol/L) (A-C) and fasting plasma glucose levels (mmol/L) (D-F) between metabolic BMI outlier groups. Results from the Malmö Offspring Study (N=3263) are shown in (A) and (D), the Malmö Diet and Cancer Study (N=3579) in (B) and (E) and the Cilento in Aging Outcomes study (N=821) in (C and F).

**Figure S5**. Differences plasma metabolite levels between correctly predicted normal weight (NW) and overestimated (OE). Results are from the Malmö Offspring Study (MOS) (N=3263) (D), the Malmö Diet and Cancer Study (N=3579) and the Cilento in Aging Outcomes study (N=821). The 13 displayed metabolites had significantly different (p<4.6e-4) levels between NW and OE in MOS, as calculated by ANOVA.

**Figure S6**. BMI (kg/m^2^) at baseline and follow-up examination in the Malmö Diet and Cancer Study (N=1416), for metabolic BMI outlier groups. Lines indicate change in BMI from baseline to follow-up examination (mean follow up time 15.6 years).

**Figure S7**. Differences in dietary intake of fruit and vegetables (g/MJ) between metabolic BMI outlier groups. Results from The Malmö Diet and Cancer Study (N=3471) in (A) and The Malmö Offspring Study (N=1526) are shown in (B).

