## Supplementary figures and images for "Metabolome-defined obesity and the risk of future diabetes and mortality"

### Figure S1

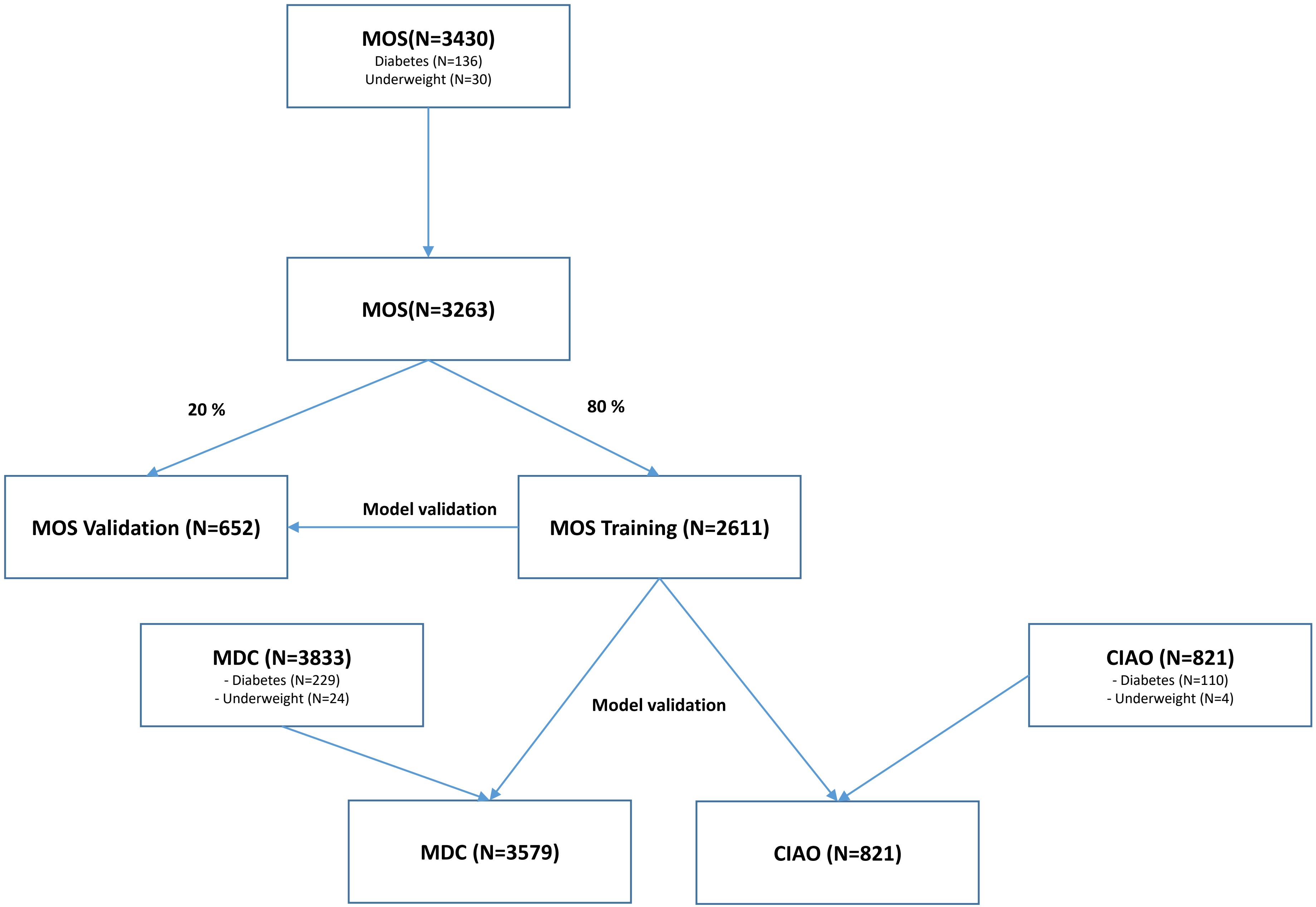

### Figure S2

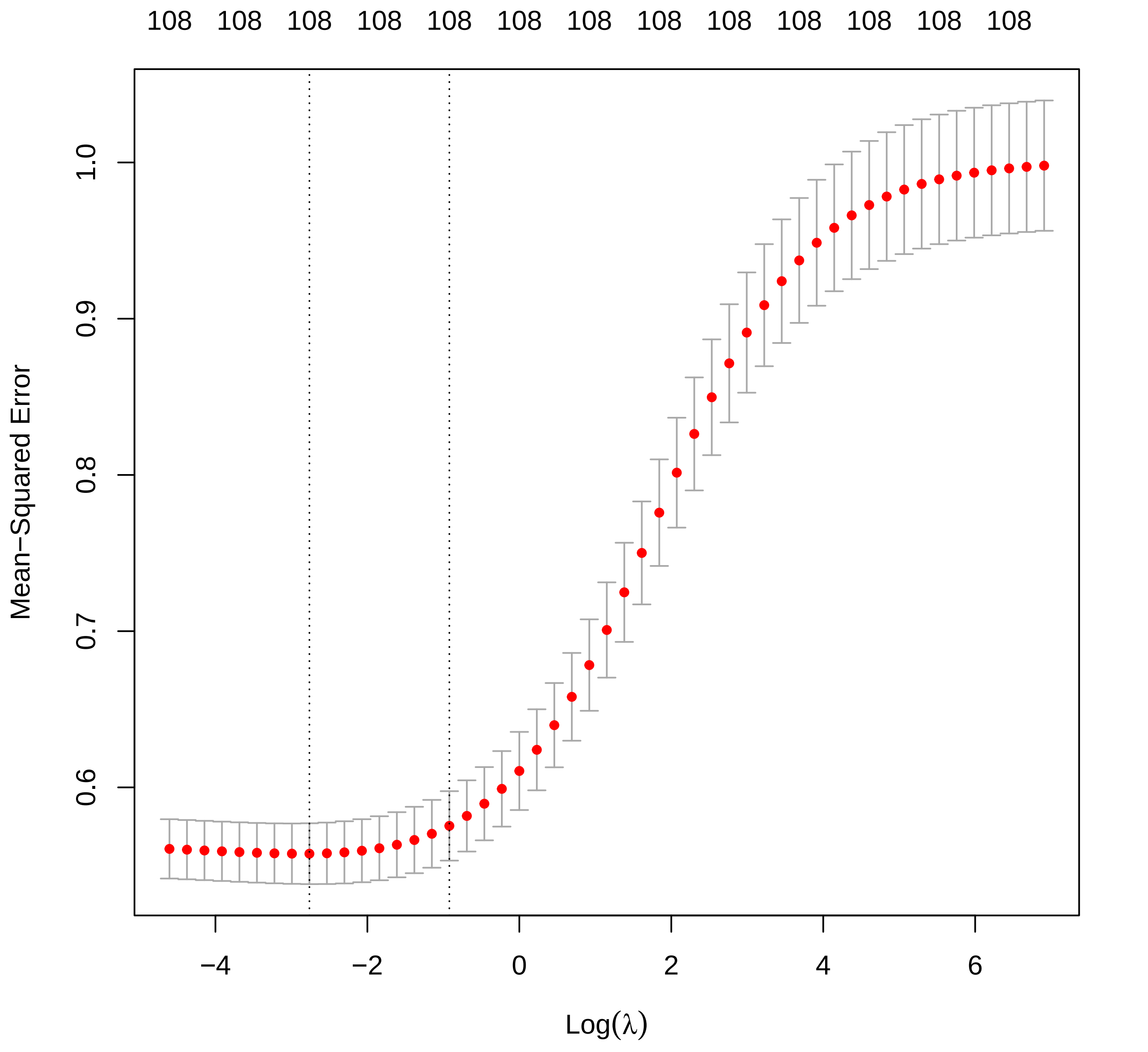

### Figure S3

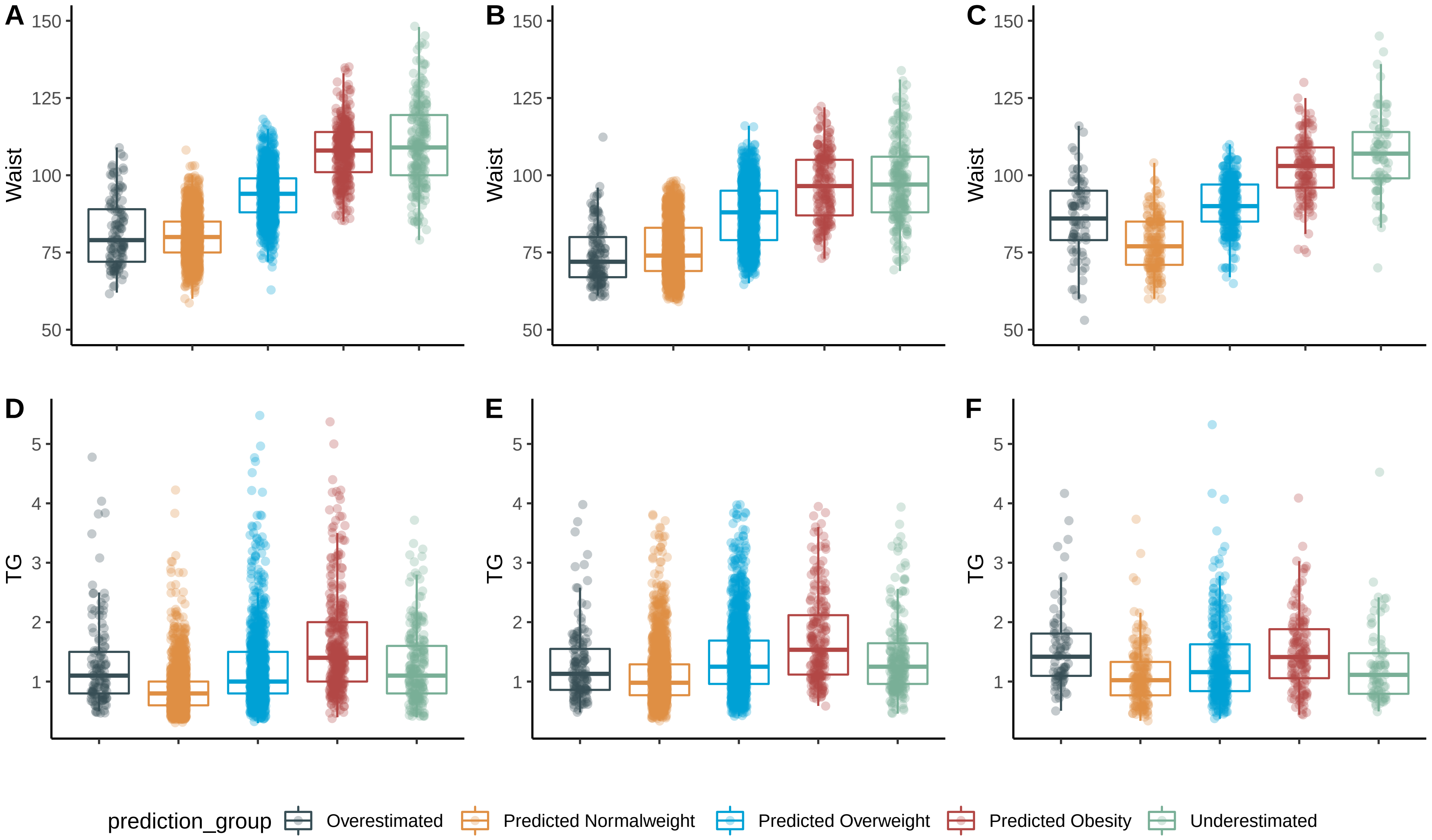

### Figure S4

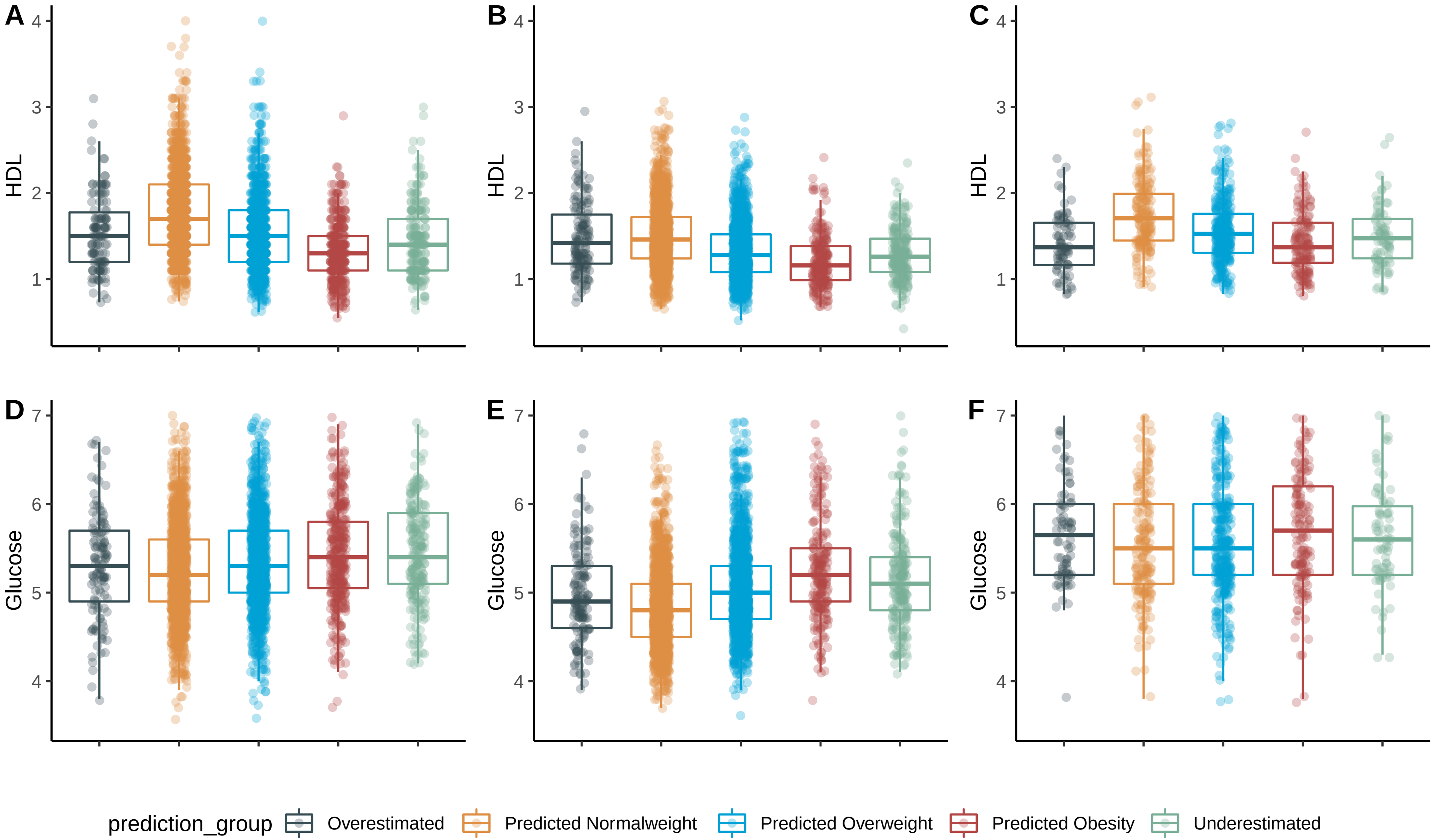

### Figure S5

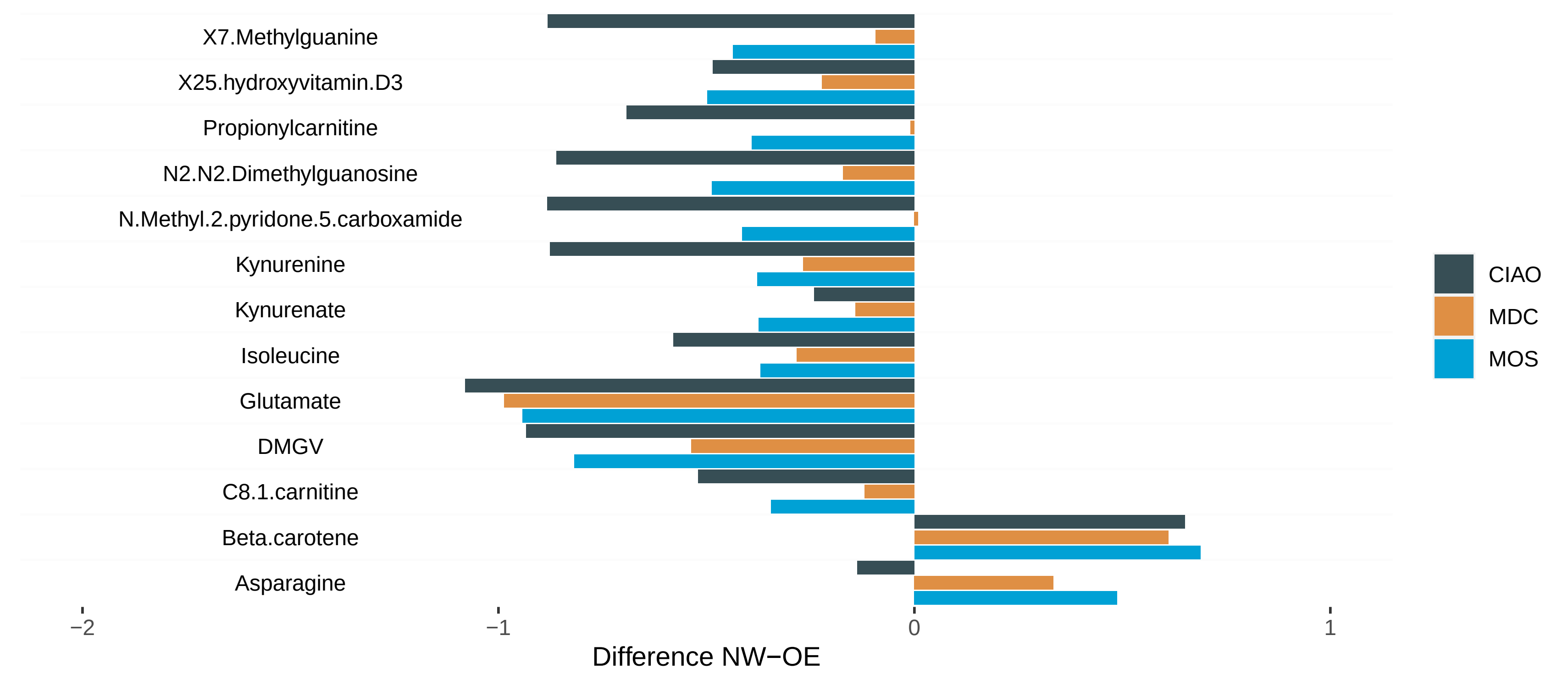

### Figure S6

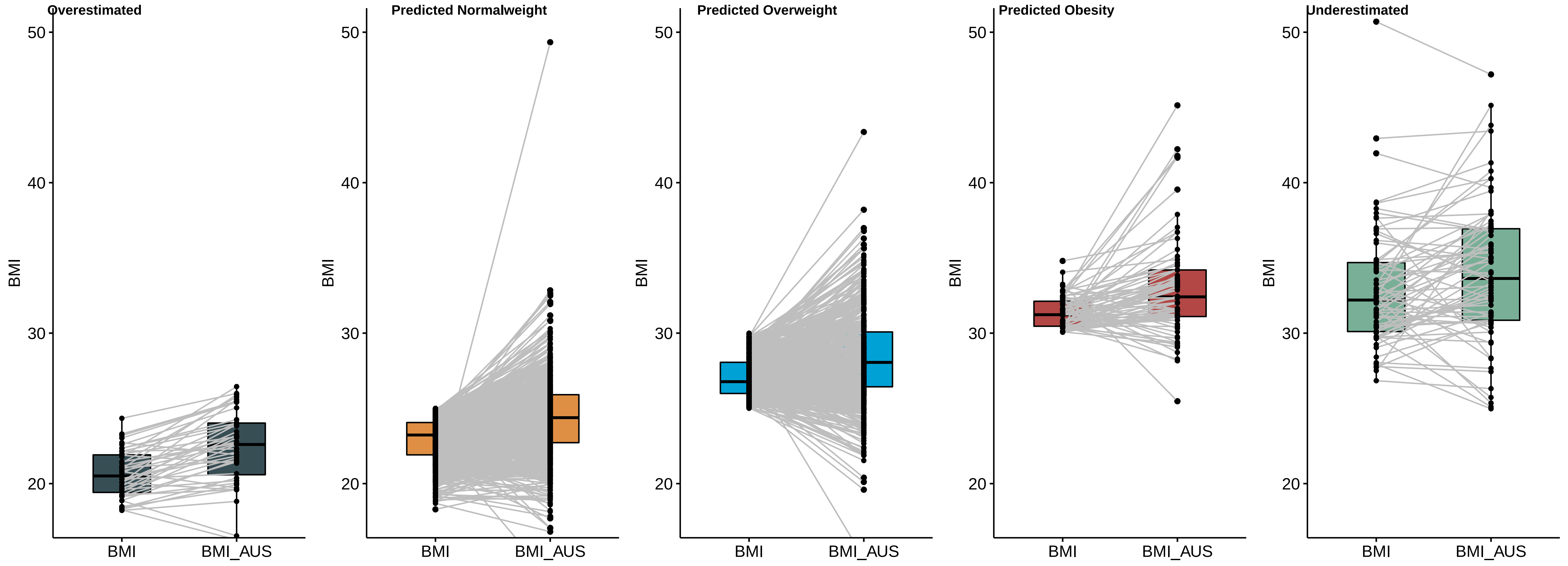

### Figure S7

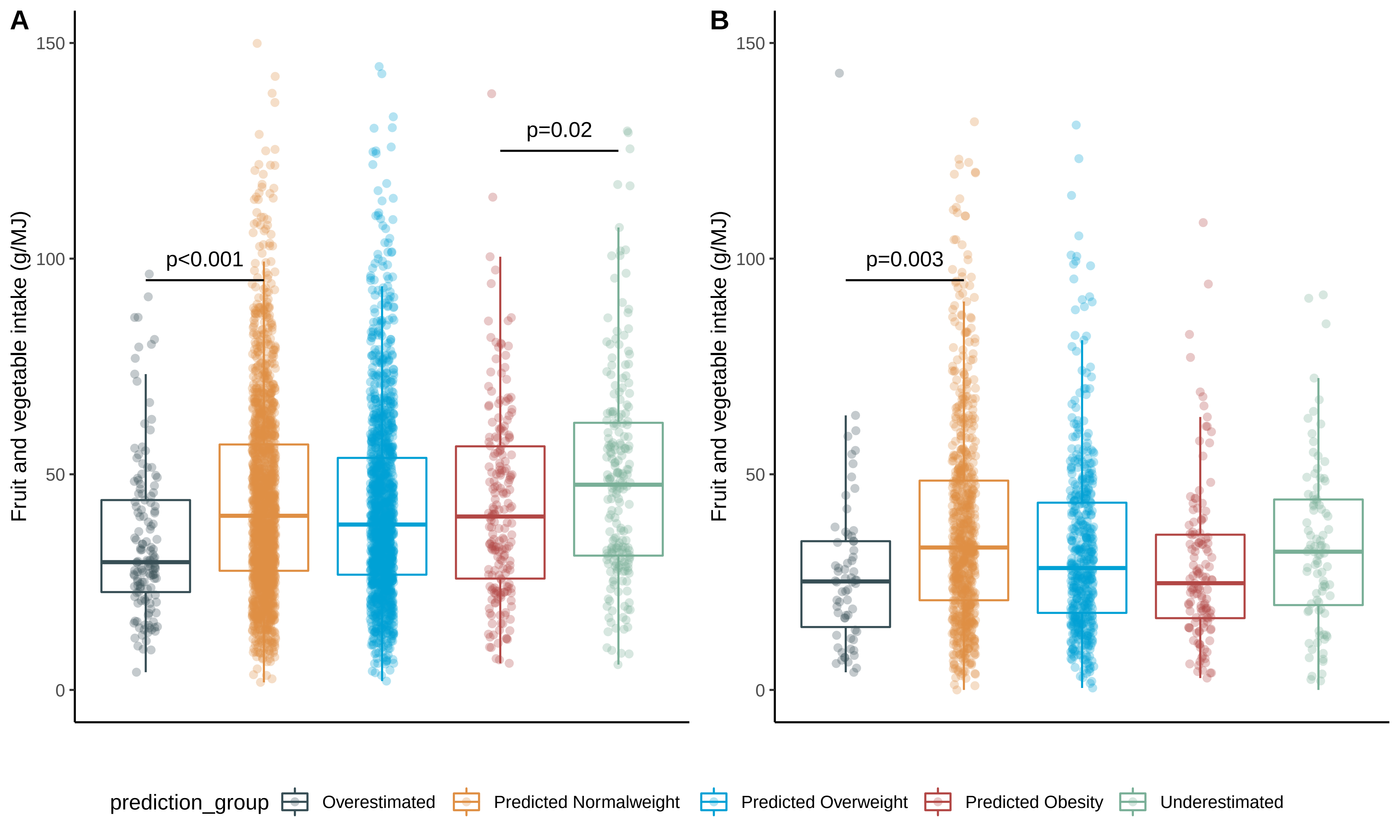
