## Supplementary material for "Metabolome-defined obesity and the risk of future diabetes and mortality": Material S1

*Biochemical measurements*

Measurements of fasting total cholesterol, HDL cholesterol, triglycerides, and glucose were made according to standard procedures at the Department of Clinical Chemistry at Malmö University Hospital. LDL cholesterol was estimated with the Friedewald equation.

*Assessment of dietary intake, physical activity and smoking*

The dietary intakes for participants from MOS were performed using a 4-day web-based food record, Riksmaten2010, which was developed by the Swedish National Food Agency^1^. In MDC, dietary intakes were assessed with a method combining a 7-day menu book, a food frequency questionnaire and a 45 minute interview^2^. Seven dietary factors related to the Swedish dietary guidelines from the Swedish Food Agency^3^ were examined: polyunsaturated fat (PUFA) (% of energy intake), saturated fat (SFA) (% of energy intake), fruits and vegetables (g/MJ), fish and shellfish (g/MJ), red meat (g/MJ), added sugar (% of energy intake) and whole-grain (MOS: g/day and MDC: portions of whole-grain meals/day). SFA and PUFA were used as markers of foods included in the guidelines that are rich in those fats, such as high-fat dairy products, high-fat meats, plant oils and nuts. In MOS, leisure-time physical activities were assessed using a questionnaire where participants were categorized as followed: 1: light exercise < 2 hours per week, 2: light exercise > 2 hours per week, 3: moderate exercise 1-2 hours per week and 4: regular exercise > 3 times per week. In MDC, the leisure-time physical activity was assessed using a questionnaire including 17 activities, which was adapted from the Minnesota Leisure Time Physical Activity Questionnaire^4^. Cigarette smoking was elicited by a self-administered questionnaire, with current cigarette smoking defined as any use within the past year.

*Metabolite normalization and annotation*

Samples were analysed in batches of 180 samples, where quality control samples were run in the beginning of each batch and every eight analytical sample, in order to condition the column and to capture analytical drift, respectively. Metabolite peak areas were integrated using Agilent Profinder B.06.00 (Agilent Technologies, Santa Clara, CA, USA). Metabolites were normalized using metabolite measurements in the quality control samples. First, a low-order nonlinear locally estimated smoothing function was fitted to the metabolite signals in the quality control samples as a function of the injection order. The α-parameter, reflecting the proportion of samples to be used when constructing the correction curve, was set to 2/3. Using this function, a correction curve for the analytical samples was interpolated, to which the metabolite measurements in the analytical samples were normalized^5^. The normalization was performed in R 3.6.1. Metabolites were annotated using synthetic standards or by matching MS/MS fragmentation with The Human Metabolome Database (HMDB)^6^ and METLIN^7^ or by matching fragment ions to putative molecule fragments. In total, 109 Identified and putatively annotated metabolites were measured. A list of reported metabolites can be found below, including corresponding mass, chromatographic retention time, molecular formula, HMDB identifier and annotation confidence^8^.

| **Metabolite** | **Mass** | **Rt** | **Formula** | **HMDB ID** | **Annotation Level** |
| --- | --- | --- | --- | --- | --- |
| Urea | 60,032 | 1,98 | CH4N2O | HMDB00294 | 1 |
| Trimethylamine-N-oxide | 75,068 | 4,28 | C3H9NO | HMDB00925 | 1 |
| Alanine | 89,048 | 7,5 | C3H7NO2 | HMDB00161 | 1 |
| Dimethylglycine | 103,063 | 6,59 | C4H9NO2 | HMDB00092 | 1 |
| 2-Aminoisobutyric acid | 103,064 | 6,9 | C4H9NO2 | HMDB01906 | 1 |
| Choline | 104,108 | 4,01 | C5H14NO | HMDB00097 | 1 |
| Serine | 105,043 | 8,56 | C3H7NO3 | HMDB00187 | 1 |
| Creatinine | 113,059 | 4,55 | C4H7N3O | HMDB00562 | 1 |
| Proline | 115,063 | 6,64 | C5H9NO2 | HMDB00162 | 1 |
| Guanidineacetate | 117,054 | 7,56 | C3H7N3O2 | HMDB00128 | 1 |
| Betaine | 118,087 | 6,14 | C5H12NO2 | HMDB00043 | 1 |
| Valine | 118,087 | 6,57 | C5H12NO2 | HMDB00883 | 1 |
| Threonine | 119,058 | 7,85 | C4H9NO3 | HMDB00167 | 1 |
| Nicotinamide | 122,048 | 1,36 | C6H6N2O | HMDB01406 | 1 |
| Taurine | 125,015 | 6,62 | C2H7NO3S | HMDB00251 | 1 |
| Pyroglutamine | 128,059 | 7,69 | C5H8N2O2 | HMDB62558 | 2 |
| Pyroglutamate | 129,043 | 2,56 | C5H7NO3 | HMDB00267 | 1 |
| N-Methylproline | 129,079 | 6,25 | C6H11NO2 | HMDB94696 | 1 |
| Pipecolate | 129,079 | 6,84 | C6H11NO2 | HMDB00716 | 1 |
| Creatine | 131,07 | 7,19 | C4H9N3O2 | HMDB00064 | 1 |
| Isoleucine | 131,095 | 6,09 | C6H13NO2 | HMDB00172 | 1 |
| Leucine | 131,095 | 5,92 | C6H13NO2 | HMDB00687 | 1 |
| Asparagine | 132,053 | 8,7 | C4H8N2O3 | HMDB00168 | 1 |
| Ornithine | 132,09 | 11,58 | C5H12N2O2 | HMDB00214 | 1 |
| Hypoxanthine | 136,039 | 3,8 | C5H4N4O | HMDB00157 | 1 |
| Trigonelline | 137,048 | 6,38 | C7H8NO2 | HMDB00875 | 1 |
| 1-Methylnicotinamide | 137,071 | 4,96 | C7H9N2O | HMDB03152 | 1 |
| Urocanate | 138,042 | 2,38 | C6H6N2O2 | HMDB00301 | 1 |
| 1,3-Dimethyluracil | 140,059 | 6,66 | C6H8N2O2 | HMDB02144 | 2 |
| Proline betaine | 143,095 | 6,21 | C7H13NO2 | HMDB04827 | 1 |
| Glutamine | 146,069 | 8,48 | C5H10N2O3 | HMDB00641 | 1 |
| Lysine | 146,106 | 11,4 | C6H14N2O2 | HMDB00182 | 1 |
| 4-Trimethylammoniobutanoic acid | 146,118 | 4,58 | C7H16NO2 | HMDB01161 | 1 |
| Glutamate | 147,053 | 8,14 | C5H9NO4 | HMDB00148 | 1 |
| Methionine | 149,051 | 6,32 | C5H11NO2S | HMDB00696 | 1 |
| Racemethionine | 149,051 | 2,23 | C5H11NO2S | HMDB33951 | 2 |
| N-Methyl-4-pyridone-3-carboxamide | 152,058 | 1,92 | C7H8N2O2 | HMDB04194 | 1 |
| Homostachydrine | 157,11 | 6,02 | C8H15NO2 | HMDB33433 | 1 |
| Methyllysine | 160,121 | 10,66 | C7H16N2O2 | HMDB02038 | 1 |
| Carnitine | 162,113 | 6,19 | C7H16NO3 | HMDB00062 | 1 |
| Methionine-S-oxide | 165,046 | 8,23 | C5H11NO3S | HMDB02005 | 1 |
| 7-Methylguanine | 165,065 | 4,77 | C6H7N5O | HMDB00897 | 1 |
| Phenylalanine | 165,079 | 5,88 | C9H11NO2 | HMDB00159 | 1 |
| 3-Methylhistidine | 169,085 | 10,82 | C7H11N3O2 | HMDB00479 | 1 |
| 1-Methylhistidine | 169,085 | 9,72 | C7H11N3O2 | HMDB00001 | 1 |
| Acetylornithine | 174,102 | 7,86 | C7H14N2O3 | HMDB03357 | 1 |
| Arginine | 174,112 | 10,92 | C6H14N4O2 | HMDB00517 | 1 |
| Citrulline | 175,095 | 9,04 | C6H13N3O3 | HMDB00904 | 1 |
| Hippurate | 179,058 | 1,14 | C9H9NO3 | HMDB00714 | 1 |
| Paraxanthine | 180,065 | 1,33 | C7H8N4O2 | HMDB01860 | 1 |
| Tyrosine | 181,074 | 6,64 | C9H11NO3 | HMDB00158 | 1 |
| Acisoga | 184,121 | 1,63 | C9H16N2O2 | HMDB61384 | 2 |
| Histidine | 188,127 | 10,56 | C6H9N3O2 | HMDB00177 | 1 |
| Homoarginine | 188,127 | 10,59 | C7H16N4O2 | HMDB00670 | 1 |
| NMMA | 188,127 | 10,2 | C7H16N4O2 | HMDB29416 | 1 |
| Kynurenate | 189,043 | 5,06 | C10H7NO3 | HMDB00715 | 1 |
| Homocitrulline | 189,111 | 8,86 | C7H15N3O3 | HMDB00679 | 1 |
| Trimethyllysine | 189,16 | 10,14 | C9H21N2O2 | HMDB01325 | 1 |
| Caffeine | 194,08 | 1 | C8H10N4O2 | HMDB01847 | 1 |
| 5-Acetylamino-6-amino-3-methyluracil | 198,075 | 4,61 | C7H10N4O3 | HMDB04400 | 1 |
| DMGV | 202,119 | 6,01 | C8H15N3O3 | HMDB0240212 | 2 |
| Asymmetric dimethylarginine | 202,143 | 9,52 | C8H18N4O2 | HMDB01539 | 1 |
| Symmetric dimethylarginine | 202,143 | 9,51 | C8H18N4O2 | HMDB03334 | 1 |
| Tryptophan | 204,09 | 5,93 | C11H12N2O2 | HMDB00929 | 1 |
| Acylcarnitine C2:0 | 204,124 | 4,45 | C9H18NO4 | HMDB00201 | 1 |
| 3-Hydroxytrimethyllysine | 205,155 | 10,8 | C9H21N2O3 | HMDB01422 | 1 |
| Kynurenine | 208,085 | 6 | C10H12N2O3 | HMDB00684 | 1 |
| Acetylarginine | 216,122 | 7,08 | C8H16N4O3 | HMDB04620 | 1 |
| Acylcarnitine C3:0 | 218,139 | 4,06 | C10H19NO4 | HMDB00824 | 1 |
| Pantothenate | 219,111 | 1,63 | C9H17NO5 | HMDB00210 | 1 |
| Ergothioneine | 229,088 | 7,35 | C9H15N3O2S | HMDB03045 | 1 |
| Acylcarnitine C4:0 | 232,154 | 3,63 | C11H21NO4 | HMDB02013 | 1 |
| Cystine | 240,024 | 13,02 | C6H12N2O4S2 | HMDB00192 | 1 |
| Tiglylcarnitine | 243,147 | 3,46 | C12H21NO4 | HMDB02366 | 2 |
| Acylcarnitine C5:0 | 245,163 | 3,23 | C12H23NO4 | HMDB00688 | 2 |
| Glycerophosphocholine | 258,11 | 8,71 | C8H20NO6P | HMDB00086 | 1 |
| Acylcarnitine C6:0 | 260,186 | 2,88 | C13H25NO4 | HMDB00756 | 2 |
| Phenylacetylglutamine | 264,11 | 1,98 | C13H16N2O4 | HMDB06344 | 1 |
| Gln.Glu | 275,117 | 10,62 | C10H17N3O6 | HMDB28796 | 2 |
| 1-Methyladenosine | 281,113 | 6,46 | C11H15N5O4 | HMDB03331 | 2 |
| Acylcarnitine C8:1 | 286,202 | 2,73 | C15H27NO4 | HMDB13324 | 2 |
| Acylcarnitine C8:0 | 288,217 | 2,49 | C15H29NO4 | HMDB00791 | 1 |
| Acylcarnitine C9:0 | 302,233 | 2,23 | C16H31NO4 | HMDB13288 | 2 |
| Acylcarnitine C8:0-OH | 304,212 | 4,06 | C15H30NO5 | | 2 |
| Acylcarnitine C10:3 | 310,202 | 2,42 | C17H28NO4 | | 2 |
| N2,N2-Dimethylguanosine | 311,123 | 4,74 | C12H17N5O5 | HMDB04824 | 1 |
| Acylcarnitine C10:2 | 312,217 | 2,44 | C17H30NO4 | | 2 |
| Acylcarnitine C10:1 | 314,232 | 2,34 | C17H31NO4 | HMDB13205 | 2 |
| Acylcarnitine C10:0 | 316,249 | 2,18 | C17H33NO4 | HMDB00651 | 2 |
| Acylcarnitine C11:1 | 328,248 | 2,28 | C18H34NO4 | | 2 |
| Acylcarnitine C11:0 | 330,263 | 2,14 | C18H35NO4 | HMDB13321 | 2 |
| Acylcarnitine C10:0-OH | 332,244 | 3,52 | C17H34NO5 | | 2 |
| Acylcarnitine C12:2 | 340,25 | 2,23 | C19H34NO4 | | 2 |
| Acylcarnitine C12:1 | 342,263 | 2,16 | C19H35NO4 | HMDB13326 | 2 |
| Acylcarnitine C12:0 | 344,279 | 2,13 | C19H37NO4 | HMDB02250 | 2 |
| Acylcarnitine C13:1 | 356,279 | 2,1 |  |  | 2 |
| Acylcarnitine C13:0 | 358,295 | 2,03 | C20H40NO4 | | 2 |
| Acylcarnitine C14:2 | 368,28 | 2,08 | C21H37NO4 | HMDB13331 | 2 |
| Acylcarnitine C14:1 | 370,294 | 2,03 | C21H39NO4 | HMDB0240588 | 2 |
| Acylcarnitine C14:0 | 372,311 | 1,94 | C21H41NO4 | HMDB05066 | 1 |
| Acylcarnitine C16:1 | 398,326 | 1,93 | C23H43NO4 | HMDB13207 | 2 |
| 25-Hydroxyvitamin D3 | 400,334 | 0,81 | C27H44O2 | HMDB03550 | 1 |
| Acylcarnitine C16:0 | 400,343 | 1,85 | C23H45NO4 | HMDB00222 | 1 |
| Acylcarnitine C18:2 | 424,346 | 1,87 | C25H46NO4 | | 2 |
| Acylcarnitine C18:1 | 426,357 | 1,86 | C25H48NO4 | | 2 |
| Acylcarnitine C18:0 | 428,372 | 1,84 | C25H49NO4 | HMDB00848 | 2 |
| Beta-carotene | 536,438 | 0,83 | C40H56 | HMDB00561 | 1 |
| Bilirubin | 584,262 | 3,91 | C33H36N4O6 | HMDB00054 | 2 |
| Urobilin | 594,342 | 1,19 | C33H46N4O6 | HMDB04160 | 2 |
